# Mid- and long-term risk of atrial fibrillation among breast cancer surgery survivors

**DOI:** 10.1101/2023.07.26.23293184

**Authors:** Yong-Moon Mark Park, Wonyoung Jung, Yohwan Yeo, Sang Hyun Park, Michael G. Fradley, Sindhu J. Malapati, Tushar Tarun, Vinay Raj, Hong Seok Lee, Tasneem Z. Naqvi, Ronda S. Henry-Tillman, Jawahar L. Mehta, Mario Schootman, Benjamin C. Amick, Kyungdo Han, Dong Wook Shin

**Affiliations:** Department of Epidemiology, Fay W. Boozman College of Public Health, University of Arkansas for Medical Sciences, Little Rock, AR; Winthrop P. Rockefeller Cancer Institute, University of Arkansas for Medical Sciences, Little Rock, AR; Department of Family Medicine, College of Medicine, Hallym University Kangdong Sacred Heart Hospital, Seoul, Republic of Korea; Department of Medicine, Sungkyunkwan University School of Medicine, Seoul, Republic of Korea; Department of Family Medicine, College of Medicine, Hallym University Dongtan Sacred Heart Hospital, Hwaseong, Republic of Korea; Department of Biostatistics, College of Medicine, The Catholic University of Korea, Seoul, Republic of Korea; Cardio-Oncology Program, Division of Cardiology, Department of Internal Medicine, University of Pennsylvania, Philadelphia, PA; Division of Medical Oncology, Department of Internal Medicine, University of Arkansas for Medical Sciences, Little Rock, AR; Division of Cardiology, Department of Medicine, University of Arkansas for Medical Sciences Little Rock, AR; Department of Biology & Department of Math and Computer Science, University of Arkansas at Pine Bluff, AR; Division of Cardiology, Sarver Heart Center, Banner University Medical Group, University of Arizona, Tucson, AZ; Division of Echocardiography, Department of Cardiovascular Medicine, Mayo Clinic, Phoenix, Arizona; Division of Breast Surgical Oncology, Department of Surgery, University of Arkansas for Medical Sciences, Little Rock, AR; Department of Medicine, College of Medicine, University of Arkansas for Medical Sciences, Little Rock, AR; Department of Statistics and Actuarial Science, Soongsil University, Seoul, Republic of Korea; Department of Family Medicine and Supportive Care Center, Samsung Medical Center, Sungkyunkwan University School of Medicine, Seoul, Republic of Korea; Department of Clinical Research Design & Evaluation, Samsung Advanced Institute for Health Science & Technology (SAIHST), Sungkyunkwan University, Seoul, Republic of Korea

**Keywords:** Atrial fibrillation, breast cancer, anthracyclines, younger breast cancer survivors

## Abstract

**Background:** The mid- and long-term risk of incident atrial fibrillation (AF) among breast cancer survivors, especially for younger women, and cancer treatment effects on the association remain unclear. We aimed to investigate the risk of AF among breast cancer survivors and evaluate the association by age group, length of follow-up, and cancer treatment.

**Methods:** Using data from the Korean Health Insurance Service database between January 2010 and December 2017, 113,232 women newly diagnosed with breast cancer (aged ≥18 years) without prior AF history who underwent breast cancer surgery were individually matched 1:5 by birth year to a sample female population without cancer (n=566,160). Sub-distribution hazard ratios (sHRs) and 95% confidence intervals (CIs) considering death as a competing risk were estimated, adjusting for sociodemographic factors and cardiovascular/non-cardiovascular comorbidities.

**Results:** During follow-up (mean [SD] follow-up, 5.1 [2.1] years), AF developed in 1,166 (1.0%) breast cancer surgery survivors at least 1 year after enrollment. Overall, breast cancer survivors had a slightly increased AF risk compared to their cancer-free counterparts (sHR 1.06; 95% CI 1.00-1.13), but the association disappeared over time. Younger breast cancer survivors (age<40 years) had more than a 2-fold increase in AF risk (sHR 2.79; 95% CI 1.98-3.94), with the association remaining similar over 5 years of follow-up. The increased risk was not observed among older breast cancer survivors, especially those aged>65 years. Breast cancer survivors who received anthracyclines had an increased risk of AF compared to those without the exposure (sHR 1.57; 95% CI 1.28-1.92) over the entire course of follow-up. The association between anthracyclines and AF incidence was also more robust in younger breast cancer survivors (sHR 1.94; 95% CI 1.40-2.69 in those aged ≤50 years). Sensitivity analyses, including further adjustments for obesity and lifestyle factors, supported the results.

**Conclusions:** Our findings suggest that younger breast cancer survivors had an elevated risk of incident AF, regardless of the length of follow-up. The use of anthracyclines may increase the mid-to-long-term AF risk among breast cancer surgery survivors.

## BACKGROUND

Individuals treated for breast cancer may be at increased risk of developing atrial fibrillation (AF), which is one of the key drivers of cardiovascular (CV) complications such as myocardial ischemia, stroke, and heart failure.[1, 2] Proposed mechanisms for increased risk of AF among breast cancer survivors include shared common risk factors such as advancing age, alcohol consumption, smoking, and obesity, as well as cancer-related proinflammatory state and cardiotoxic cancer treatment.[1–3] Due to increased early detection and improved survival following breast cancer treatment,[4] mid- to long-term CV complications have become a major concern of breast cancer survivors.[2]

The evidence regarding the risk of AF among breast cancer survivors has been primarily derived from studies limited by relatively small numbers of patients evaluated over a short period.[5] Additionally, most studies have focused on women over 65 years,[6, 7] or have been limited by the short-term follow-up (e.g., <3 years),[6, 8] small numbers of incident AF events,[9] use of prevalent breast cancer cases,[8, 10] early-stage breast cancer,[11] and failure to consider competing risks (e.g., death)[8, 12, 13] and comprehensive cancer treatment effects.[12–14] Consequently, the risk of AF among breast cancer survivors is unknown.

Breast cancer and CV events in women vary with age, particularly before and after menopause onset.[2] For instance, compared to those without cancer, women aged <40 years with breast cancer are at more than a 3-fold increased risk of developing CV complications,[15] and women diagnosed with breast cancer before age 50 years are at a higher risk of developing treatment-related CV complications.[16] However, few studies have investigated whether AF risk differs between younger and older breast cancer survivors,[8] particularly in Asian women, whose breast cancer incidence peaks in the mid to late 40s, which is remarkably different from that observed in Western countries.[17] Using the Korean National Health Insurance Service (NHIS) database, we evaluated incident AF among breast cancer survivors compared with women without cancer, particularly investigating the mid- to long-term risk of AF and the impact of cancer treatment on this association by the age at the time of breast cancer diagnosis.

## METHODS

### Data Source and Study Setting

The NHIS is a public health insurance program providing mandatory universal coverage to 97% of the South Korean population. The NHIS database comprises sociodemographic and claims-based healthcare information such as medical procedures, diagnoses, prescriptions, outpatient visits, and hospital admission.[18, 19] The NHIS also operates biennial standardized health screening examinations,[20] which include past medical history, lifestyle behaviors (smoking, drinking, and physical activity), anthropometric measurements, and laboratory tests for nonemployees aged ≥40 years and employees regardless of age. The NHIS database has been validated for epidemiological and clinical research.[21, 22]

This study was approved by the Samsung Medical Center (Seoul, South Korea; SMC 2020-03-108) Institutional Review Board. All information used for analyses was anonymized and de-identified; therefore, informed consent was not required. The database is open to all researchers whose study protocols are approved by the official review committee.

### Study Population

We identified 129,548 women newly diagnosed with invasive breast cancer who had undergone breast cancer surgery between January 1, 2010, and December 31, 2017, based on both International Classification of Diseases, Tenth Revision (ICD-10) codes (C50) and cancer-specific insurance claim code (V193 code). This V code was used to ensure the accuracy of breast cancer diagnosis because it is a reimbursement code representing biopsy-confirmed cancers in South Korea.[23] We excluded those who did not undergo breast cancer surgery to avoid late-stage or aggressive breast cancer cases. Additionally, we excluded those who had a history of any cancer (n=12,043), were younger than 18 years old (n=17), or had a prior history of AF (n=2,148). We further excluded person-time within the first year of follow-up to reduce the bias related to undetected AF present at baseline and the acute effect of breast cancer treatments on AF risk[1] (n=2,108). The resulting 113,232 breast cancer survivors were matched 1:5 to women with no history of cancer or AF (n=566,160) from the general population based on birth year at baseline. Data recorded through December 31, 2020, were included in our analyses (**Figure 1**).

**Figure 1.**
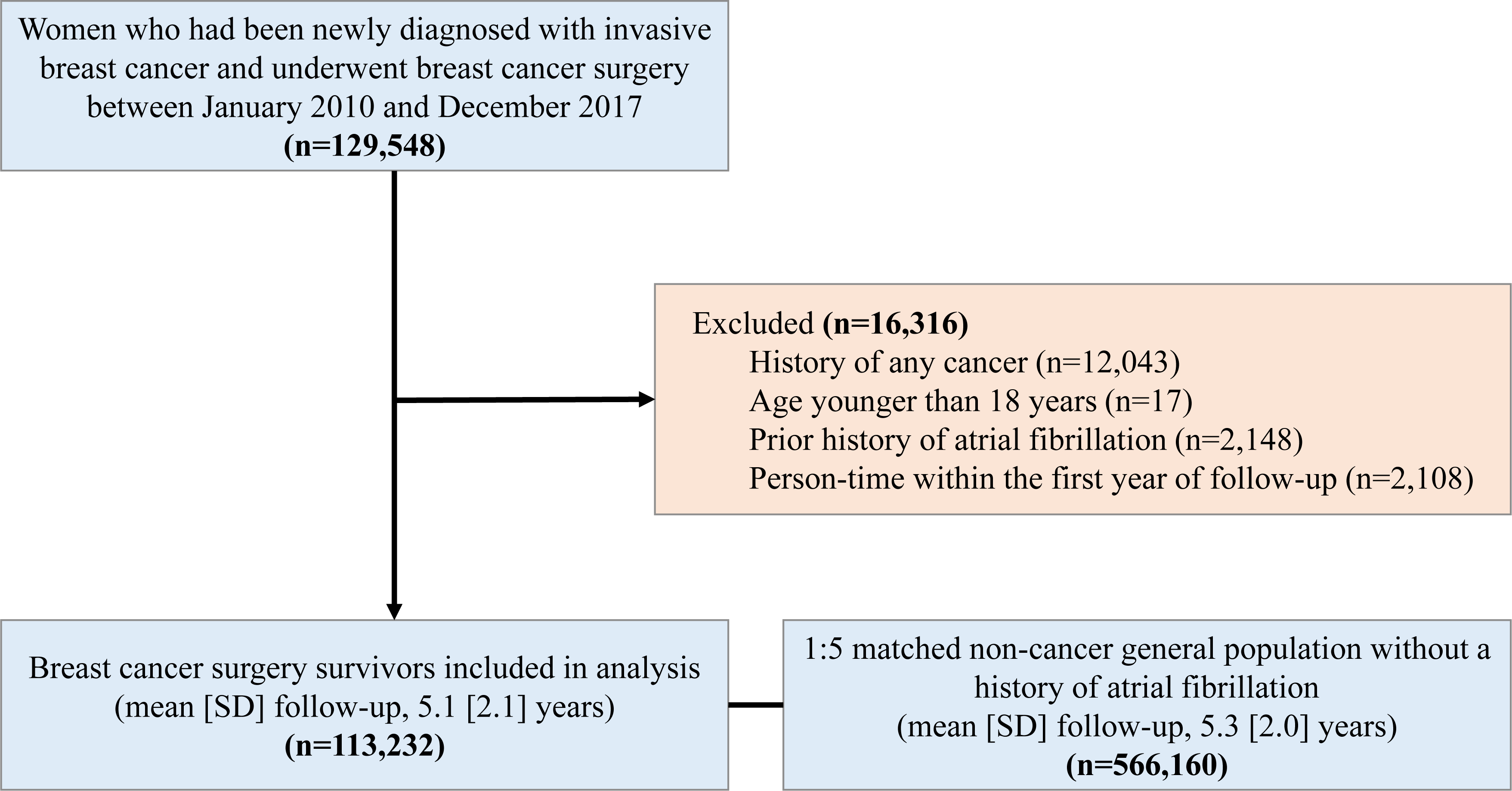
Participant flow diagram. After excluding participants meeting exclusion criteria, 113,232 subjects were included in the final analysis.

### Measurements

The primary outcome was incident AF, based on ICD-10 codes I48.0–I48.4, and I48.9. AF diagnosis using ICD-10 codes in the NHIS has been shown to be 94.1% accurate.[24] We defined individuals with AF as those who had a discharge diagnosis or visited an outpatient clinic more than twice to exclude those with transient AF and improve diagnostic accuracy.[25]

Breast cancer treatment information was based on the claims data within 1 year after the breast cancer diagnosis.[23, 26] Chemotherapy was defined as at least 1 treatment cycle of a chemotherapeutic agent (anthracyclines [epirubicin or doxorubicin], cyclophosphamide, and taxane-based regimens [docetaxel or paclitaxel]). Due to the reimbursement policy in the NHIS, most patients administered taxane-based regimens tended to take anthracyclines in the adjuvant setting, indicating that those given taxanes could also be included in a category of those who took anthracyclines.[26] Targeted therapy was defined as at least one treatment cycle of Herceptin (trastuzumab). Endocrine therapy was defined as treatment with tamoxifen or aromatase inhibitors (anastrozole, exemestane, and letrozole), categorizing the use of a specific regimen based on the initial prescription when there was a subsequent switching between medicines. Radiation therapy was defined as at least one local or regional treatment.

Comorbid medical conditions were assessed using past medical history data and clinical and pharmacy ICD-10 codes. Comorbidities included hypertension, type 2 diabetes, dyslipidemia, coronary heart disease (CHD), congestive heart failure, stroke, chronic kidney disease, and chronic obstructive pulmonary disease (COPD). Comorbidities of participants were identified based on laboratory measures, claims, and prescription information prior to the index date as follows: hypertension (ICD-10 codes [I10.x-I13.x and I15.x], or being on antihypertensive medication or having blood pressure ≥ 140/90 mmHg), diabetes mellitus (DM) (ICD-10 codes [E11.x-E14.x] with antidiabetic medications, or a fasting glucose level ≥ 126 mg/dL), dyslipidemia (ICD-10 code E78.x with lipid-lowering medication, or total cholesterol level ≥ 240 mg/dL), coronary heart disease (ICD-10 codes [I21.x-I22.x] during hospitalization), congestive heart failure (ICD codes [I50.x] for the first hospitalization), stroke (ICD-10 codes [I63.x-I64.x] during hospitalization, with claims for brain magnetic resonance imaging or brain computed tomography), chronic kidney disease (the glomerular filtration rate of <60mL/min/1.73 m2 as estimated by the Modification of Diet in Renal Disease equation), and chronic obstructive pulmonary disease (ICD-10 codes [J43.x-J44.x]). ICD-10 codes were also applied if recorded in at least two outpatient visits for coronary heart disease and stroke. In addition, the Charlson Comorbidity Index (CCI) was calculated based on ICD-10 codes. Income level was based on monthly health insurance premiums. Low-income status was defined as being in the lowest quartile of monthly health insurance premiums or being enrolled in the Medical Aid program. The geographic area of residence was dichotomized by rural and urban areas using the primary local authority districts (shi/gun/gu).

### Statistical Analysis

The baseline characteristics are presented as means with standard deviations or numbers with percentages. Participant data were collected until the date of AF diagnosis, death, or end of the study, whichever occurred first. Person-time was calculated starting at 1 year after baseline. The crude AF incidence rate was assessed by dividing the number of events by the total number of person-years of follow-up presented as per 1,000 person-years. Competing risk survival statistics considering death as a competing risk were used to calculate the cumulative incidence of AF.[27] The AF incidence between women with and without breast cancer was compared using the Gray-K test.[28]

The Fine-Gray proportional sub-distribution hazards model was used to estimate sub-distribution hazard ratios (sHRs) and 95% confidence intervals (CIs) for AF incidence with death as a competing risk.[29] The proportional hazards assumption was assessed using Schoenfeld’s residuals, and no specific departure was observed. In addition to a crude model (Model 1), potential confounders were identified a priori based on a literature review; these included age at baseline (continuous variable), household income, and residential location, which were incorporated into Model 2 to account for sociodemographic characteristics. Model 3 further adjusted for the presence or absence of hypertension, type 2 diabetes, dyslipidemia, CHD, congestive heart failure, stroke, chronic kidney disease, and COPD to account for comorbidities. To examine AF risk by cancer treatment modalities among breast cancer survivors, we further adjusted for the use of anthracyclines, taxanes, trastuzumab, endocrine treatment, and radiation treatment in Model 4. All analyses were stratified by age categories (18–39, 40–50, 51–65, or ≥66 years). Landmark analyses were performed using 2 landmark points (3 and 5 years after breast cancer diagnosis) to estimate AF risk in an unbiased way in individuals who were event-free at the landmark time and examine the long-term effects of breast cancer and its treatments on AF incidence.[30]

As a sensitivity analysis, we included person-time within the first year of follow-up to capture any short-term CV consequences. In addition, using a subset of data comprising participants in the general health screening examination, we conducted sensitivity analyses among 72,560 women with and 292,468 without breast cancer to account for body mass index, smoking, alcohol consumption, and physical activity (see **Supplemental Methods** for measurement details). A potential effect modification by income status, residential location, and other comorbidities was evaluated through stratified analysis and interaction testing using a likelihood ratio test. All statistical analyses were performed using SAS version 9.4 (SAS Institute Inc., Cary, NC, USA). The *P* values provided are two-sided, with the level of significance at 0.05.

## RESULTS

### Baseline characteristics

Baseline characteristics are shown in **Table 1**, stratified by the presence or absence of breast cancer. The mean study population age was 51.6 years, and 10.8% of participants were less than 40 years old. Individuals surgically treated for breast cancer (hereafter referred to as breast cancer survivors) were more likely to have hypertension, type 2 diabetes, dyslipidemia, CHD, congestive heart failure, chronic kidney disease, and COPD than women without cancer. Among breast cancer survivors, the proportions of use in each treatment option were 61.0% for chemotherapy, 15.0% for target therapy, 61.8% for endocrine treatment, and 69.8% for radiation therapy. Breast cancer survivors were less likely to have low incomes or live in rural areas. Overall, the prevalence of CV risk factors and comorbidities was higher in breast cancer survivors than in women without cancer in each age group **(Supplemental Table 1)**.

**Table 1.**
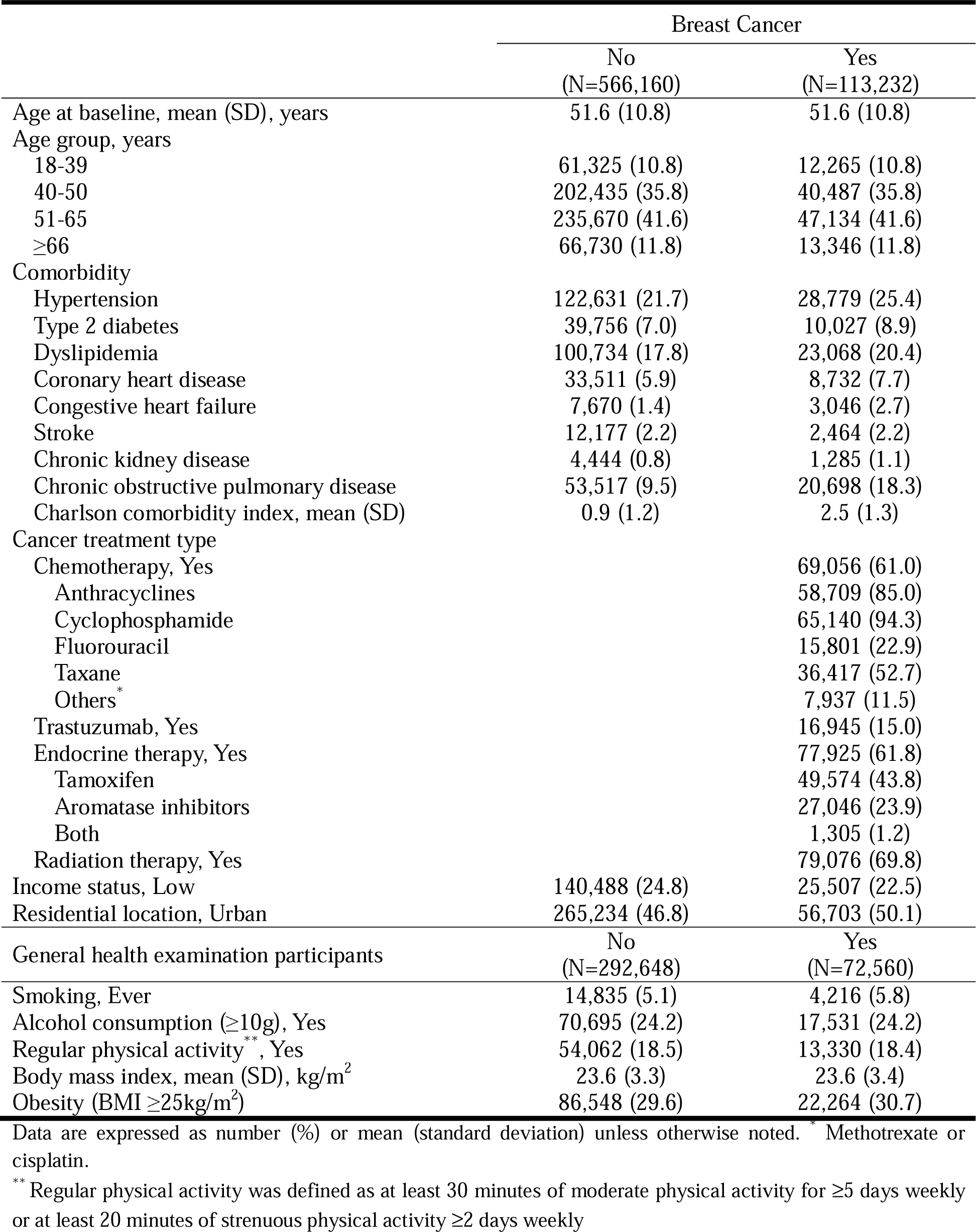
Baseline characteristics of the study population.

|  | Breast Cancer |  |
| --- | --- | --- |
|  | No<br>(N=566,160) | Yes<br>(N=113,232) |
| Age at baseline, mean (SD), years | 51.6 (10.8) | 51.6 (10.8) |
| Age group, years |  |  |
| 18-39 | 61,325 (10.8) | 12,265 (10.8) |
| 40-50 | 202,435 (35.8) | 40,487 (35.8) |
| 51-65 | 235,670 (41.6) | 47,134 (41.6) |
| ≥66 | 66,730 (11.8) | 13,346 (11.8) |
| Comorbidity |  |  |
| Hypertension | 122,631 (21.7) | 28,779 (25.4) |
| Type 2 diabetes | 39,756 (7.0) | 10,027 (8.9) |
| Dyslipidemia | 100,734 (17.8) | 23,068 (20.4) |
| Coronary heart disease | 33,511 (5.9) | 8,732 (7.7) |
| Congestive heart failure | 7,670 (1.4) | 3,046 (2.7) |
| Stroke | 12,177 (2.2) | 2,464 (2.2) |
| Chronic kidney disease | 4,444 (0.8) | 1,285 (1.1) |
| Chronic obstructive pulmonary disease | 53,517 (9.5) | 20,698 (18.3) |
| Charlson comorbidity index, mean (SD) | 0.9 (1.2) | 2.5 (1.3) |
| Cancer treatment type |  |  |
| Chemotherapy, Yes |  | 69,056 (61.0) |
| Anthracyclines |  | 58,709 (85.0) |
| Cyclophosphamide |  | 65,140 (94.3) |
| Fluorouracil |  | 15,801 (22.9) |
| Taxane |  | 36,417 (52.7) |
| Others* |  | 7,937 (11.5) |
| Trastuzumab, Yes |  | 16,945 (15.0) |
| Endocrine therapy, Yes |  | 77,925 (61.8) |
| Tamoxifen |  | 49,574 (43.8) |
| Aromatase inhibitors |  | 27,046 (23.9) |
| Both |  | 1,305 (1.2) |
| Radiation therapy, Yes |  | 79,076 (69.8) |
| Income status, Low | 140,488 (24.8) | 25,507 (22.5) |
| Residential location, Urban | 265,234 (46.8) | 56,703 (50.1) |
| General health examination participants | No<br>(N=292,648) | Yes<br>(N=72,560) |
| Smoking, Ever | 14,835 (5.1) | 4,216 (5.8) |
| Alcohol consumption (≥10g), Yes | 70,695 (24.2) | 17,531 (24.2) |
| Regular physical activity**, Yes | 54,062 (18.5) | 13,330 (18.4) |
| Body mass index, mean (SD), kg/m<sup>2</sup> | 23.6 (3.3) | 23.6 (3.4) |
| Obesity (BMI ≥25kg/m<sup>2</sup>) | 86,548 (29.6) | 22,264 (30.7) |

Data are expressed as number (%) or mean (standard deviation) unless otherwise noted. \* Methotrexate or cisplatin.
\*\* Regular physical activity was defined as at least 30 minutes of moderate physical activity for ≥5 days weekly or at least 20 minutes of strenuous physical activity ≥2 days weekly

### Risk of AF in breast cancer survivors compared to the general population by age group

Risks of AF by age group are shown in **Figure 2** and **Table 2**. A total of 1,166 (1.0%) incident AF cases were identified at least 1 year after baseline during a mean (SD) of 5.1 (2.1) years of follow-up in breast cancer survivors. AF risk was 6% higher in breast cancer survivors than those without cancer (Model 3: sHR 1.06, 95% CI 1.00-1.13) after adjusting for potential confounders. Associations differed by age group (*P* for interaction <0.001). Younger women (18–39 years) with breast cancer exhibited a 2.79-fold increased AF risk (sHR 2.79, 95% CI 1.98-3.94), whereas older women (≥66 years) with breast cancer exhibited a decreased AF risk (sHR 0.90, 95% CI 0.81-0.99), compared with those without cancer. In the landmark analyses, AF risk was attenuated over time, and there was no overall association with AF over 5 years after breast cancer diagnosis. However, younger women with breast cancer exhibited a sustained 2-fold increased risk in those aged <40 years and at least a 30% increased AF risk in those aged ≤50 years during follow-up **(Supplemental Table 2)**. In contrast, the inverse association tended to persist in those aged >65 years (*P* for interaction for age group in both landmark analyses <0.05).

**Figure 2.**
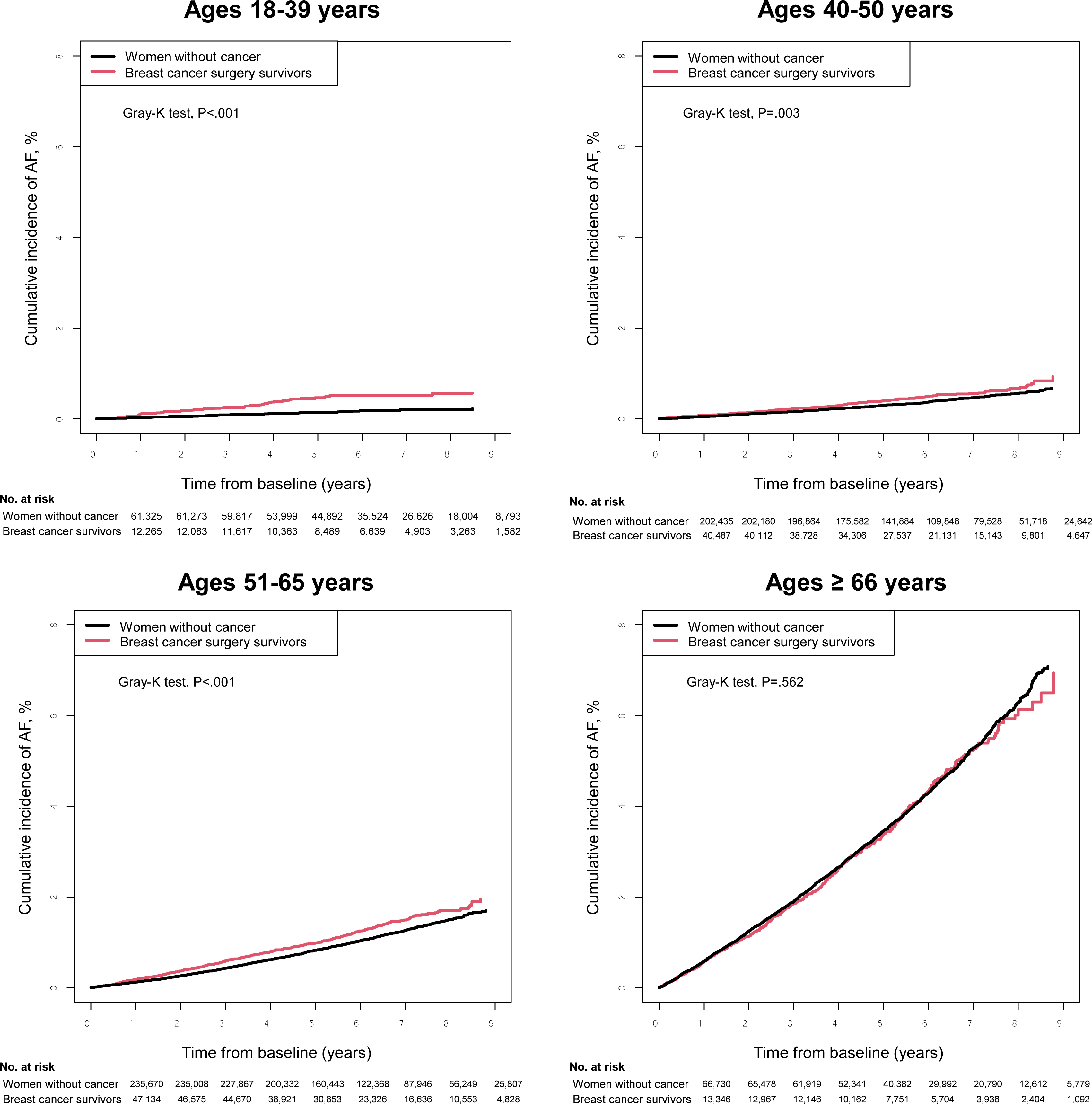
Cumulative incidence of Atrial Fibrillation (AF) by age group. Cumulative incidence function plots for atrial fibrillation (AF) in breast cancer surgery survivors display that breast cancer surgery survivors had a consistently higher incidence of AF compared to their age-matched noncancer female general population in those aged 18-39 (Figure 2a, P<0.001), aged 40-50 (Figure 2b, P=0.003), and aged 51-65 (Figure 2c, P<0.001).

**Table 2.** Adjusted sub-distribution hazard ratios for developing atrial fibrillation in breast cancer surgery survivors compared to the noncancer general population by age categories.

|  | Age group |  | Subjects (N) | Case (n) | IR per 1,000 person-years | Model 1 (Crude)<br>sHR (95% CI) | Model 2<br>sHR (95% CI) | Model 3<br>sHR (95% CI) |
| --- | --- | --- | --- | --- | --- | --- | --- | --- |
| Main analysis | All ages | Noncancer | 566,160 | 5,271 | 1.76 | 1(Ref.) | 1(Ref.) | 1(Ref.) |
|  |  | Breast cancer | 113,232 | 1,166 | 2.01 | 1.14 (1.07-1.22) | 1.15 (1.08-1.23) | 1.06 (1.00-1.13) |
|  | 18-39 | Noncancer | 61,325 | 91 | 0.27 | 1(Ref.) | 1(Ref.) | 1(Ref.) |
|  |  | Breast cancer | 12,265 | 51 | 0.78 | 2.94 (2.09-4.15) | 2.95 (2.10-4.16) | 2.79 (1.98-3.94) |
|  | 40-50 | Noncancer | 202,435 | 684 | 0.63 | 1(Ref.) | 1(Ref.) | 1(Ref.) |
|  |  | Breast cancer | 40,487 | 171 | 0.81 | 1.28 (1.09-1.52) | 1.29 (1.09-1.53) | 1.22 (1.03-1.45) |
|  | 51-65 | Noncancer | 235,670 | 2,123 | 1.71 | 1(Ref.) | 1(Ref.) | 1(Ref.) |
|  |  | Breast cancer | 47,134 | 497 | 2.06 | 1.21 (1.10-1.33) | 1.22 (1.10-1.34) | 1.13 (1.03-1.25) |
|  | ≥66 | Noncancer | 66,730 | 2,373 | 7.31 | 1(Ref.) | 1(Ref.) | 1(Ref.) |
|  |  | Breast cancer | 13,346 | 447 | 7.08 | 0.97 (0.88-1.07) | 0.98 (0.89-1.09) | 0.90 (0.81-0.99) |
| P for interaction |  |  |  |  |  | <0.001 | <0.001 | <0.001 |
| 3-year landmark analysis | All ages | Noncancer | 546,453 | 3,609 | 1.94 | 1(Ref.) | 1(Ref.) | 1(Ref.) |
|  |  | Breast cancer | 107,153 | 777 | 2.17 | 1.12 (1.04-1.21) | 1.13 (1.05-1.22) | 1.05 (0.97-1.14) |
|  | 18-39 | Noncancer | 59,817 | 60 | 0.28 | 1(Ref.) | 1(Ref.) | 1(Ref.) |
|  |  | Breast cancer | 11,617 | 31 | 0.76 | 2.75 (1.78-4.24) | 2.76 (1.79-4.26) | 2.62 (1.70-4.05) |
|  | 40-50 | Noncancer | 196,863 | 472 | 0.69 | 1(Ref.) | 1(Ref.) | 1(Ref.) |
|  |  | Breast cancer | 38,727 | 117 | 0.89 | 1.28 (1.05-1.57) | 1.29 (1.06-1.58) | 1.23 (1.00-1.50) |
|  | 51-65 | Noncancer | 227,856 | 1,516 | 1.97 | 1(Ref.) | 1(Ref.) | 1(Ref.) |
|  |  | Breast cancer | 44,665 | 328 | 2.22 | 1.13 (1.00-1.27) | 1.14 (1.01-1.28) | 1.06 (0.94-1.20) |
|  | ≥66 | Noncancer | 61,917 | 1,561 | 8.04 | 1(Ref.) | 1(Ref.) | 1(Ref.) |
|  |  | Breast cancer | 12,144 | 301 | 8.06 | 1.00 (0.89-1.14) | 1.02 (0.90-1.15) | 0.93 (0.82-1.05) |
| P for interaction |  |  |  |  |  | <0.001 | <0.001 | <0.001 |
| 5-year landmark analysis | All ages | Noncancer | 546,453 | 1,918 | 2.11 | 1(Ref.) | 1(Ref.) | 1(Ref.) |
|  |  | Breast cancer | 107,153 | 388 | 2.25 | 1.07 (0.96-1.19) | 1.08 (0.96-1.19) | 1.00 (0.90-1.12) |
|  | 18-39 | Noncancer | 59,817 | 28 | 0.25 | 1(Ref.) | 1(Ref.) | 1(Ref.) |
|  |  | Breast cancer | 11,617 | 12 | 0.59 | 2.32 (1.18-4.56) | 2.33 (1.18-4.57) | 2.22 (1.13-4.37) |
|  | 40-50 | Noncancer | 196,863 | 265 | 0.79 | 1(Ref.) | 1(Ref.) | 1(Ref.) |
|  |  | Breast cancer | 38,727 | 65 | 1.01 | 1.28 (0.98-1.68) | 1.29 (0.98-1.69) | 1.23 (0.94-1.62) |
|  | 51-65 | Noncancer | 227,856 | 812 | 2.18 | 1(Ref.) | 1(Ref.) | 1(Ref.) |
|  |  | Breast cancer | 44,665 | 163 | 2.31 | 1.06 (0.90-1.25) | 1.06 (0.90-1.26) | 1.00 (0.84-1.18) |
|  | ≥66 | Noncancer | 61,917 | 813 | 9.08 | 1(Ref.) | 1(Ref.) | 1(Ref.) |
|  |  | Breast cancer | 12,144 | 148 | 8.70 | 0.96 (0.80-1.14) | 0.96 (0.81-1.15) | 0.89 (0.74-1.06) |
| P for interaction |  |  |  |  |  | 0.041 | 0.041 | 0.024 |

IR, incidence rate; PYs, person-years; sHR, sub-distribution hazard ratio; CI, confidence interval.
Model 2: adjusted for age, income status, and residential location.

### Risk of AF by treatment modalities

Risks of AF by cancer treatment modalities among breast cancer survivors are shown in **Table 3**. Individuals who received anthracyclines had a 57% higher AF risk compared with those who did not (Model 4: sHR 1.57, 95% CI 1.28-1.92), whereas tamoxifen users had a 19% lower AF risk compared with those who did not receive endocrine treatment (sHR 0.81, 95% CI 0.70-0.95). Taxane-based chemotherapy was associated with increased AF risk (Model 3: sHR 1.20, 95% CI 1.06-1.35), but the association became nonsignificant after further adjusting for other cancer treatment modalities (Model 4: sHR 0.84, 95% CI 0.69-1.02). Other cancer therapies, including trastuzumab, aromatase inhibitors, and radiation treatment, were not associated with AF incidence. In the landmark analyses, the use of anthracyclines showed persistent increased AF risk over the follow-up period. In contrast, the inverse association between tamoxifen use and AF risk tended to disappear.

**Table 3.** Adjusted sub-distribution hazard ratios for developing atrial fibrillation by cancer treatment type among breast cancer surgery survivors.

|  | Treatment type |  | Subjects (N) | Case (n) | IR per 1,000 PYs | Model 1 (Crude) sHR (95% CI) | Model 2 sHR (95% CI) | Model 3 sHR (95% CI) | Model 4 sHR (95% CI) |
| --- | --- | --- | --- | --- | --- | --- | --- | --- | --- |
| **Main analysis** | **Anthracyclines** | No | 54,523 | 591 | 2.14 | 1(Ref.) | 1(Ref.) | 1(Ref.) | 1(Ref.) |
|  |  | Yes | 58,709 | 575 | 1.89 | 0.88 (0.79-0.98) | 1.39 (1.23-1.58) | 1.39 (1.23-1.57) | 1.57 (1.28-1.92) |
|  | **Taxane** | No | 48,092 | 530 | 2.17 | 1(Ref.) | 1(Ref.) | 1(Ref.) | 1(Ref.) |
|  |  | Yes | 65,140 | 636 | 1.89 | 0.87 (0.77-0.97) | 1.22 (1.08-1.37) | 1.20 (1.06-1.35) | 0.84 (0.69-1.02) |
|  | **Trastuzumab** | No | 96,287 | 996 | 2.00 | 1(Ref.) | 1(Ref.) | 1(Ref.) | 1(Ref.) |
|  |  | Yes | 16,945 | 170 | 2.04 | 1.03 (0.87-1.21) | 1.15 (0.97-1.35) | 1.11 (0.94-1.30) | 0.96 (0.80-1.14) |
|  | **Endocrine therapy** | No | 35,307 | 386 | 2.19 | 1(Ref.) | 1(Ref.) | 1(Ref.) | 1(Ref.) |
|  |  | Tamoxifen | 49,574 | 294 | 1.12 | 0.51 (0.44-0.59) | 0.79 (0.68-0.92) | 0.78 (0.67-0.92) | 0.81 (0.70-0.95) |
|  |  | AIs | 27,046 | 471 | 3.46 | 1.58 (1.38-1.81) | 1.01 (0.88-1.15) | 1.00 (0.87-1.15) | 1.00 (0.87-1.15) |
|  |  | Both | 1,305 | 15 | 2.23 | 1.02 (0.61-1.70) | 0.70 (0.42-1.17) | 0.66 (0.40-1.11) | 0.68 (0.41-1.14) |
|  | **Radiation therapy** | No | 34,156 | 433 | 2.52 | 1(Ref.) | 1(Ref.) | 1(Ref.) | 1(Ref.) |
|  |  | Yes | 79,076 | 733 | 1.79 | 0.71 (0.63-0.80) | 1.01 (0.89-1.14) | 1.02 (0.90-1.16) | 0.98 (0.87-1.12) |
| **3-year landmark analysis** | **Anthracyclines** | No | 52,036 | 390 | 2.32 | 1(Ref.) | 1(Ref.) | 1(Ref.) | 1(Ref.) |
|  |  | Yes | 55,117 | 387 | 2.04 | 0.88 (0.76-1.01) | 1.41 (1.22-1.64) | 1.40 (1.21-1.63) | 1.48 (1.16-1.90) |
|  | **Taxane** | No | 45,695 | 345 | 2.31 | 1(Ref.) | 1(Ref.) | 1(Ref.) | 1(Ref.) |
|  |  | Yes | 61,458 | 432 | 2.07 | 0.90 (0.78-1.03) | 1.27 (1.10-1.48) | 1.25 (1.08-1.45) | 0.91 (0.72-1.16) |
|  | **Trastuzumab** | No | 91,190 | 663 | 2.15 | 1(Ref.) | 1(Ref.) | 1(Ref.) | 1(Ref.) |
|  |  | Yes | 15,963 | 114 | 2.28 | 1.07 (0.88-1.30) | 1.19 (0.98-1.46) | 1.16(0.95-1.41) | 0.99 (0.80-1.23) |
|  | **Endocrine therapy** | No | 32,452 | 252 | 2.34 | 1(Ref.) | 1(Ref.) | 1(Ref.) | 1(Ref.) |
|  |  | Tamoxifen | 47,839 | 208 | 1.27 | 0.54 (0.45-0.65) | 0.85 (0.70-1.03) | 0.85 (0.70-1.02) | 0.89 (0.74-1.08) |
|  |  | AIs | 25,633 | 308 | 3.72 | 1.59 (1.35-1.88) | 0.99 (0.84-1.18) | 0.99(0.83-1.17) | 0.99 (0.83-1.17) |
|  |  | Both | 1,229 | 9 | 2.16 | 0.92 (0.47-1.79) | 0.62 (0.32-1.21) | 0.59(0.30-1.14) | 0.61 (0.31-1.18) |
|  | **Radiation therapy** | No | 31,842 | 283 | 2.70 | 1(Ref.) | 1(Ref.) | 1(Ref.) | 1(Ref.) |
|  |  | Yes | 75,311 | 494 | 1.95 | 0.72 (0.62-0.84) | 1.04 (0.89-1.21) | 1.05 (0.90-1.22) | 1.01 (0.87-1.18) |
| **5-year landmark analysis** | **Anthracyclines** | No | 34,664 | 183 | 2.32 | 1(Ref.) | 1(Ref.) | 1(Ref.) | 1(Ref.) |
|  |  | Yes | 39,965 | 205 | 2.20 | 0.95 (0.78-1.16) | 1.57 (1.27-1.94) | 1.56 (1.26-1.93) | 1.56 (1.10-2.21) |
|  | **Taxane** | No | 31,154 | 160 | 2.28 | 1(Ref.) | 1(Ref.) | 1(Ref.) | 1(Ref.) |
|  |  | Yes | 43,475 | 228 | 2.24 | 0.98 (0.80-1.20) | 1.43 (1.16-1.76) | 1.40 (1.14-1.73) | 1.02 (0.72-1.43) |
|  | **Trastuzumab** | No | 64,089 | 332 | 2.22 | 1(Ref.) | 1(Ref.) | 1(Ref.) | 1(Ref.) |
|  |  | Yes | 10,540 | 56 | 2.45 | 1.11 (0.84-1.47) | 1.24 (0.93-1.65) | 1.20 (0.90-1.59) | 1.01 (0.75-1.37) |
|  | **Endocrine therapy** | No | 22,325 | 115 | 2.23 | 1(Ref.) | 1(Ref.) | 1(Ref.) | 1(Ref.) |
|  |  | Tamoxifen | 34,191 | 116 | 1.46 | 0.65 (0.50-0.84) | 1.04 (0.80-1.35) | 1.04 (0.80-1.35) | 1.13 (0.86-1.48) |
|  |  | AIs | 17,253 | 152 | 3.89 | 1.74 (1.37-2.22) | 1.07 (0.83-1.36) | 1.06 (0.83-1.36) | 1.08 (0.84-1.38) |
|  |  | Both | 860 | 5 | 2.49 | NA | NA | NA | NA |
|  | **Radiation therapy** | No | 21,464 | 144 | 2.86 | 1(Ref.) | 1(Ref.) | 1(Ref.) | 1(Ref.) |
|  |  | Yes | 53,165 | 244 | 2.00 | 0.70 (0.57-0.86) | 1.02 (0.82-1.26) | 1.03 (0.83-1.28) | 0.98 (0.79-1.22) |

IR, incidence rate; PYs, person-years; sHR, sub-distribution hazard ratio; CI, confidence interval; AIs, aromatase inhibitors; NA, Not applicable.

Risks of AF according to cancer treatment modalities and age group among breast cancer survivors are shown in **Table 4**. The use of anthracyclines was associated with increased AF risk in all age groups, with a stronger association among younger women aged ≤50 years. The inverse association between tamoxifen use and AF risk also tended to be clearer among younger women aged ≤50 years **(Supplemental Table 3)**. In the landmark analyses, the use of anthracyclines tended to be consistently associated with increased AF risk in each age group over the follow-up. In contrast, the inverse association between tamoxifen use and AF risk was no longer significant **(Supplemental Table 4)**.

**Table 4.**
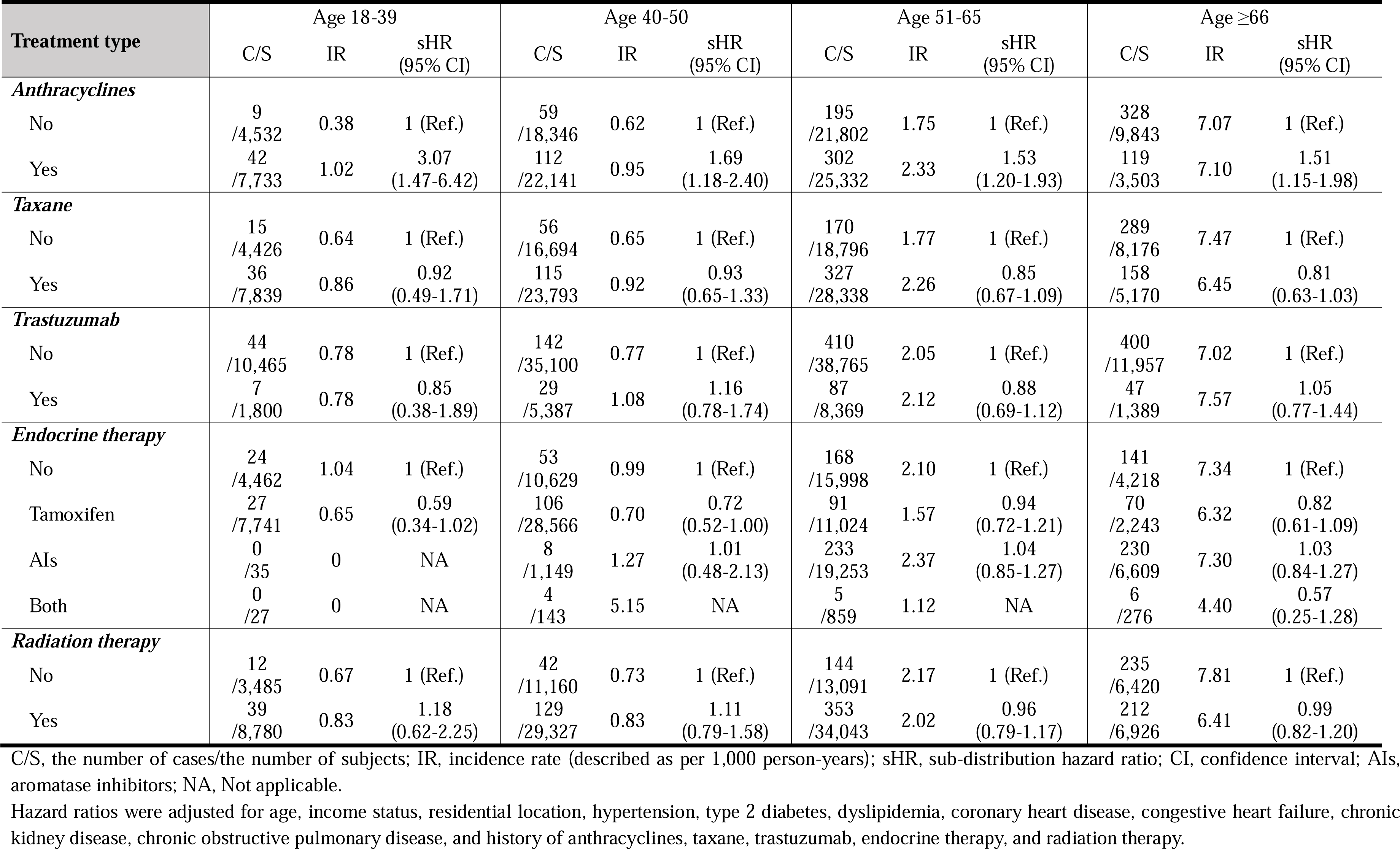
Adjusted sub-distribution hazard ratios for developing atrial fibrillation by cancer treatment type among breast cancer surgery survivors by age categories.

### Stratified and sensitivity analyses

In stratified analyses, increased AF risk was observed only for those without CHD (*P* for interaction=0.04) or those without COPD (*P* for interaction=0.06) among women with breast cancer of all ages **(Supplemental Table 5)**. When we included person-time within the first year of follow-up, the associations tended to be stronger, particularly in younger breast cancer survivors (aged ≤50 years) **(Supplemental Table 6)**. In contrast, meaningful changes were not observed in risks of AF by cancer treatment modalities among breast cancer survivors **(Supplemental Table 7).** The results were not materially changed in analyses limited to participants in the NHIS health examinations where body mass index, smoking, alcohol consumption, and physical activity were further adjusted for **(Supplemental Tables 8 & 9)**.

## DISCUSSION

In this nationwide population-based cohort study, younger breast cancer survivors had increased AF risk compared to an age-matched sample of females with no cancer history from the general population; the strength of this association remained persistent over 5 years of follow-up. Breast cancer survivors aged <40 years had a more than 2-fold increased AF risk during the follow-up; this elevated risk was not observed in older breast cancer survivors, particularly those aged >65 years. Among breast cancer survivors, those treated with anthracyclines had a more pronounced AF risk than those not exposed to this chemotherapeutic. The association was strongest in breast cancer survivors aged ≤50 years and remained persistent over the follow-up period.

Newly developed AF in individuals with cancer may have an adverse effect on prognosis. In a study using the US Surveillance, Epidemiology, and End Results Medicare registry data, older women aged >65 years with breast cancer who developed incident AF within the first 30 days of breast cancer diagnosis were 3 times more likely to die of a CV condition within 1 year compared with those without incident AF.[6] In a Taiwanese retrospective cohort study, individuals with cancer who developed new-onset AF had an increased risk of thromboembolism and heart failure, although all-cause mortality was not elevated.[31] In contrast, new-onset AF in those with lymphoma was positively associated with both acute heart failure and all-cause mortality.[32] In addition, there was a higher risk of bleeding in patients with cancer and AF than in those without AF.[33]

While we initially observed an overall positive association between breast cancer and the risk of developing AF, it was marginal and became null over the follow-up period. These results are similar to previous studies, which were unable to show a strong association between breast cancer and incident AF compared to other types of cancer.[13, 14] The strength of the association with the risk of incident AF also decreased over time, with the highest risk occurring in the first year of diagnosis,[6, 7, 11, 13, 14] which was also observed in our study. This transient, increased risk of developing AF shortly after breast cancer diagnosis might be due to surgical and/or medical cancer therapy, autonomic nervous system imbalance, pre-existing chronic inflammatory state, cancer-related comorbidities, or the combination of these conditions.[1] To assess the mid- to long-term risk of AF among breast cancer survivors, we excluded AF incidence in the first year of follow-up to mitigate the effects of these potential confounding factors.

In our study, younger survivors, particularly those <40 years, showed a stronger association with incident AF than older breast cancer survivors. The increased risk remained persistent over 5 years of the follow-up after adjusting for cardiometabolic comorbidities. A previous study reported that breast cancer survivors <60 years had a stronger association with incident AF than older breast cancer survivors.[8] It is unclear why the association differs by age group. In our stratified analyses, among those without CV risk factors or CVD at baseline, breast cancer survivors had a higher AF risk compared to the general population. This suggests that AF incidence between older breast cancer survivors and the general population might not be different due to the increased prevalence of CV risk factors and comorbid CVD as women age. However, the prevalence of CV risk factors and comorbidities in women aged >50 years was still higher in breast cancer survivors compared to the general population in our study. The other speculation is that younger patients receive more intensive, cardiotoxic treatments than older patients.[8] As such, regular cardiac surveillance or monitoring for AF may be warranted, although further research is needed to establish a specific prevention strategy.[34]

We observed a positive association between the use of anthracyclines and subsequent increased AF risk after accounting for the use of other cancer treatment modalities. After adjusting for other cancer treatment modalities, there was no significant association between the use of taxanes and AF occurrence, suggesting that the observed unadjusted association was likely driven by the concomitant use of anthracyclines.[26] This is consistent with prior findings from a recent study of the World Health Organization dataset VigiBase of more than 130 countries, which reported that those who developed AF were more likely to use anthracyclines, adjusting for the use of other anticancer medications.[35] In contrast, our findings differ from those of previous studies from the US[6] and Canada,[11] where anthracycline use was not associated with increased AF risk, possibly because of different study population characteristics such as racial/ethnic differences.[36] However, racial/ethnic difference in cardiotoxicity is understudied and further studies are warranted to confirm the observed finding.[37] An inverse association between tamoxifen use and AF risk among younger breast cancer survivors aged ≤50 years tended to diminish gradually with increasing follow-up duration. This finding suggests that the benefit of tamoxifen use may be limited to reduced risk of early CV outcomes in breast cancer survivors.[38]

Given the limitations of claims data, we cannot provide clinical details, including cancer stage, cardiac imaging, pathological results, surgery types, and chemotherapy dosage. In addition, misclassification and unmeasured confounding may exist as a result of limitations inherent to claims data. However, we investigated the mid- to long-term risk of AF after a breast cancer diagnosis and its surgical/medical treatments using data from a large sample size of 113,232 women newly diagnosed with breast cancer. While other non–cancer-related factors may have contributed to the development of AF, we did perform various sensitivity analyses, including further adjustment for obesity and lifestyle characteristics to minimize bias.

## CONCLUSIONS

In conclusion, young breast cancer surgery survivors, specifically those aged <40 years and those treated with anthracyclines, may be at a higher mid- to long-term risk of AF. Our findings underscore the need for increased awareness of AF risk in this population.

## LIST OF ABBREVIATIONS

AF: atrial fibrillation
CHD: coronary heart disease
CI: confidence interval
COPD: chronic obstructive pulmonary disease
CV: cardiovascular
DM: diabetes mellitus
NHIS: National Health Insurance Service
SD: standard deviation
sHR: sub-distribution hazard ratio

## Data Availability Statement

The data will be made available upon request and approval of a proposal by the National Health Insurance Service Database.

## Ethics Approval and Consent to Participate

This study was approved by the Samsung Medical Center (Seoul, South Korea; SMC 2020-03-108) Institutional Review Board. All information used for analyses was anonymized and de-identified; therefore, informed consent was not required.

## Consent for publication

Not applicable

## Acknowledgement

None

## FIGURE LEGENDS

**Figure 3.** Graphical Abstract. The graphical abstract illustrates the objectives, methods, and findings of this study.

