## Supplemental Tables for "Mid- and long-term risk of atrial fibrillation among breast cancer surgery survivors"

**Supplemental Table 1.** Comparison of baseline cardiovascular risk factors and comorbidities by age group

|  | Aged 18-39 | | | Aged 18-50 | | | Aged ≥ 51 | | |
| --- | --- | --- | --- | --- | --- | --- | --- | --- | --- |
|  | Noncancer  (n=61,325) | Breast cancer  (n=12,265) | P value | Noncancer  (n=263,760) | Breast cancer  (n=52,752) | P value | Noncancer  (n=302,400) | Breast cancer  (n=60,480) | P value |
| Income status, low | 12,698 (20.7) | 2,239 (18.3) | <.001 | 62,961 (23.9) | 11,162 (21.2) | <.001 | 77,527 (25.6) | 14,345 (23.7) | <.001 |
| Residential location, Urban | 29,358 (47.9) | 6,061 (49.4) | <.001 | 124,104 (47.1) | 25,930 (49.2) | <.001 | 141,130 (46.7) | 30,773 (50.9) | <.001 |
| Hypertension | 1,166 (1.9) | 522 (4.3) | <.001 | 17,950 (6.8) | 4,846 (9.2) | <.001 | 104,681 (34.6) | 23,933 (39.6) | <.001 |
| Type 2 diabetes | 504 (0.8) | 165 (1.4) | <.001 | 5,730 (2.2) | 1,527 (2.9) | <.001 | 34,026 (11.3) | 8,500 (14.1) | <.001 |
| Dyslipidemia | 832 (1.4) | 366 (3.0) | <.001 | 13,406 (5.1) | 3,521 (6.7) | <.001 | 87,328 (28.9) | 19,547 (32.3) | <.001 |
| Coronary heart disease | 653 (1.1) | 320 (2.6) | <.001 | 5,054 (1.9) | 1,816 (3.4) | <.001 | 28,457 (9.4) | 6,916 (11.4) | <.001 |
| Congestive heart failure | 123 (0.2) | 191 (1.6) | <.001 | 11,009 (4.2) | 832 (1.6) | <.001 | 6,661 (2.2) | 2,214 (3.7) | <.001 |
| Chronic kidney disease | 170 (0.3) | 51 (0.4) | 0.104 | 998 (0.4) | 324 (0.6) | <.001 | 3,446 (1.1) | 961 (1.6) | <.001 |
| COPD | 3,619 (5.9) | 1,804 (14.7) | <.001 | 16,356 (6.2) | 7,382 (14.0) | <.001 | 37,161 (12.3) | 13,316 (22.0) | <.001 |

COPD; chronic obstructive pulmonary disease.

**Supplemental Table 2.** Adjusted sub-distribution hazard ratios for developing atrial fibrillation in breast cancer surgery survivors compared to the noncancer general population (aged 18-50 years)

|  | | Subjects (N) | Case (n) | IR per 1,000 person-years | Model 1 (Crude)  HR (95% CI) | Model 2  sHR (95% CI) | Model 3  sHR (95% CI) |
| --- | --- | --- | --- | --- | --- | --- | --- |
| ***Main analysis*** | Noncancer | 263,760 | 775 | 0.54 | 1(Ref.) | 1(Ref.) | 1(Ref.) |
|  | Breast cancer | 52,752 | 222 | 0.80 | 1.48 (1.27-1.72) | 1.48 (1.28-1.72) | 1.40 (1.21-1.63) |
| ***3-year landmark analysis*** | Noncancer | 256,680 | 532 | 0.59 | 1(Ref.) | 1(Ref.) | 1(Ref.) |
|  | Breast cancer | 50,344 | 148 | 0.86 | 1.45 (1.21-1.74) | 1.45 (1.21-1.74) | 1.38 (1.15-1.66) |
| ***5-year landmark analysis*** | Noncancer | 256,680 | 293 | 0.66 | 1(Ref.) | 1(Ref.) | 1(Ref.) |
|  | Breast cancer | 50,344 | 77 | 0.91 | 1.39 (1.08-1.78) | 1.39 (1.08-1.78) | 1.33 (1.03-1.71) |

IR, incidence rate; HR, hazard ratio; sHR, sub-distribution hazard ratio; CI, confidence interval.

Model 2: adjusted for age, income status, and residential location.

Model 3: adjusted for Model 2 + hypertension, type 2 diabetes, dyslipidemia, coronary heart disease, congestive heart failure, chronic kidney disease, and chronic obstructive pulmonary disease.

**Supplemental Table 3.** Adjusted sub-distribution hazard ratios for developing atrial fibrillation by cancer treatment type among breast cancer surgery survivors aged 18-50 years

|  | | ***Main analysis*** | | | | ***3-year landmark analysis*** | | | | ***5-year landmark analysis*** | | | |
| --- | --- | --- | --- | --- | --- | --- | --- | --- | --- | --- | --- | --- | --- |
|  | | Subjects (N) | Case (n) | IR | sHR (95% CI) | Subjects (N) | Case (n) | IR | sHR (95% CI) | Subjects (N) | Case (n) | IR | sHR (95% CI) |
| ***Anthracycline*** | No | 22,878 | 68 | 0.57 | 1 (Ref.) | 22,117 | 45 | 0.62 | 1(Ref.) | 15,059 | 21 | 0.61 | 1(Ref.) |
|  | Yes | 29,874 | 154 | 0.97 | 1.94 (1.40-2.69) | 28,227 | 103 | 1.03 | 1.79 (1.20-2.68) | 20,967 | 56 | 1.12 | 1.86 (1.05-3.29) |
| ***Taxane*** | No | 21,120 | 71 | 0.65 | 1 (Ref.) | 20,274 | 44 | 0.65 | 1(Ref.) | 14,161 | 19 | 0.59 | 1(Ref.) |
|  | Yes | 31,632 | 151 | 0.90 | 0.94 (0.68-1.31) | 30,070 | 104 | 0.99 | 1.08 (0.72-1.62) | 21,865 | 58 | 1.11 | 1.31 (0.72-2.37) |
| ***Trastuzumab*** | No | 45,565 | 186 | 0.77 | 1 (Ref.) | 43,531 | 125 | 0.83 | 1(Ref.) | 31,416 | 63 | 0.85 | 1(Ref.) |
|  | Yes | 7,187 | 36 | 1.00 | 1.09 (0.76-1.57) | 6,813 | 23 | 1.06 | 1.08 (0.69-1.69) | 4,610 | 14 | 1.38 | 1.37 (0.76-2.45) |
| ***Endocrine treatment*** | No | 15,091 | 77 | 1.01 | 1 (Ref.) | 13,903 | 45 | 0.95 | 1(Ref.) | 9,785 | 24 | 1.04 | 1(Ref.) |
|  | Tamoxifen | 36,307 | 133 | 0.69 | 0.66 (0.50-0.87) | 35,151 | 95 | 0.79 | 0.79 (0.56-1.13) | 25,303 | 50 | 0.85 | 0.78 (0.48-1.27) |
|  | AIs | 1,184 | 8 | 1.24 | 0.85 (0.41-1.76) | 1,137 | 7 | 1.71 | 1.21 (0.55-2.70) | 832 | 3 | 1.43 | 0.88 (0.27-2.93) |
|  | Both | 170 | 4 | 4.52 | NA | 153 | 1 | 1.79 | 1.41 (0.19-10.21) | 106 | 0 | 0 | NA |
| ***Radiation treatment*** | No | 14,645 | 54 | 0.72 | 1 (Ref.) | 13,831 | 35 | 0.76 | 1(Ref.) | 9,455 | 13 | 0.58 | 1(Ref.) |
|  | Yes | 38,107 | 168 | 0.83 | 1.13 (0.83-1.54) | 36,513 | 113 | 0.89 | 1.15 (0.79-1.68) | 26,571 | 64 | 1.03 | 1.69 (0.93-3.07) |

IR, incidence rate (described as per 1,000 person-years); HR, hazard ratio; sHR, sub-distribution hazard ratio; CI, confidence interval; AIs, aromatase inhibitors.

Hazard ratios were adjusted for income, area of residence, hypertension, diabetes mellitus, dyslipidemia, coronary heart disease, congestive heart failure, chronic kidney disease, chronic obstructive pulmonary disease, history of anthracyclines, taxane, trastuzumab, endocrine therapy, and radiation therapy.

**Supplemental Table 4.** Landmark analyses at the 3- and 5-year landmark points for adjusted sub-distribution hazard ratios for developing atrial fibrillation by age categories and cancer treatment type among breast cancer surgery survivors

|  | | ***3-year landmark analysis*** | | | | ***5-year landmark analysis*** | | | | |
| --- | --- | --- | --- | --- | --- | --- | --- | --- | --- | --- |
| ***Age 18-39*** | | Subjects (N) | Case (n) | IR | sHR (95% CI) | Subjects (N) | Case (n) | IR | | sHR (95% CI) |
| ***Anthracycline*** | No | 4,365 | 6 | 0.40 | 1(Ref.) | 3,044 | 4 | | 0.55 | 1(Ref.) |
|  | Yes | 7,252 | 25 | 0.96 | 2.50 (1.00-6.23) | 5,445 | 8 | | 0.61 | 1.07 (0.31-3.68) |
| ***Taxane*** | No | 4,215 | 9 | 0.61 | 1(Ref.) | 3,025 | 6 | | 0.83 | 1(Ref.) |
|  | Yes | 7,402 | 22 | 0.84 | 0.98 (0.44,2.18) | 5,464 | 6 | | 0.45 | 0.39 (0.12-1.24) |
| ***Trastuzumab*** | No | 9,927 | 27 | 0.76 | 1(Ref.) | 7,309 | 12 | | 0.67 | 1(Ref.) |
|  | Yes | 1,690 | 4 | 0.73 | 0.823(0.288,2.356) | 1,180 | 0 | | 0 | NA |
| ***Endocrine treatment*** | No | 4,080 | 12 | 0.83 | 1(Ref.) | 2,965 | 6 | | 0.83 | 1(Ref.) |
|  | Tamoxifen | 7,485 | 19 | 0.72 | 0.82 (0.40-1.68) | 5,497 | 6 | | 0.46 | 0.52 (0.17-1.61) |
|  | AIs | 32 | 0 | 0 | NA | 18 | 0 | | 0 | NA |
|  | Both | 20 | 0 | 0 | NA | 9 | 0 | | 0 | NA |
| ***Radiation treatment*** | No | 3,257 | 5 | 0.45 | 1(Ref.) | 2238 | 2 | | 0.37 | 1(Ref.) |
|  | Yes | 8,360 | 26 | 0.87 | 1.84(0.71,4.79) | 6251 | 10 | | 0.66 | 1.71 (0.37-7.79) |
| ***Age 40-50*** | | Subjects (N) | Case (n) | IR | sHR (95% CI) | Subjects (N) | Case (n) | IR | | sHR (95% CI) |
| ***Anthracycline*** | No | 17,752 | 39 | 0.67 | 1(Ref.) | 12,015 | 17 | | 0.62 | 1(Ref.) |
|  | Yes | 20,975 | 78 | 1.06 | 1.62 (1.05-2.49) | 15,522 | 48 | | 1.30 | 2.02 (1.09-3.74) |
| ***Taxane*** | No | 16,059 | 35 | 0.66 | 1(Ref.) | 11,136 | 13 | | 0.52 | 1(Ref.) |
|  | Yes | 22,668 | 82 | 1.04 | 1.11 (0.71-1.73) | 16,401 | 52 | | 1.33 | 1.80 (0.92-3.53) |
| ***Trastuzumab*** | No | 33,604 | 98 | 0.85 | 1(Ref.) | 24,107 | 51 | | 0.90 | 1(Ref.) |
|  | Yes | 5,123 | 19 | 1.17 | 1.15 (0.70-1.89) | 3,430 | 14 | | 1.86 | 1.70 (0.94-3.09) |
| ***Endocrine treatment*** | No | 9,823 | 33 | 1.00 | 1(Ref.) | 6,820 | 18 | | 1.14 | 1(Ref.) |
|  | Tamoxifen | 27,666 | 76 | 0.80 | 0.82 (0.54-1.23) | 19,806 | 44 | | 0.96 | 0.86 (0.50-1.50) |
|  | AIs | 1,105 | 7 | 1.74 | 1.38 (0.61-3.12) | 814 | 3 | | 1.45 | 0.98 (0.29-3.34) |
|  | Both | 133 | 1 | 2.00 | 1.66 (0.23-12.11) | 97 | 0 | | 0 | NA |
| ***Radiation treatment*** | No | 10,574 | 30 | 0.85 | 1(Ref.) | 7,217 | 11 | | 0.65 | 1(Ref.) |
|  | Yes | 28,153 | 87 | 0.90 | 1.03 (0.68-1.56) | 20,320 | 54 | | 1.15 | 1.68 (0.88-3.22) |
| ***Age 51-65*** | | Subjects (N) | Case (n) | IR | sHR (95% CI) | Subjects (N) | Case (n) | IR | | sHR (95% CI) |
| ***Anthracycline*** | No | 20,936 | 120 | 1.76 | 1(Ref.) | 13,965 | 53 | | 1.66 | 1(Ref.) |
|  | Yes | 23,729 | 208 | 2.60 | 1.59 (1.18-2.13) | 16,887 | 110 | | 2.84 | 1.75 (1.15-2.66) |
| ***Taxane*** | No | 17,976 | 103 | 1.75 | 1(Ref.) | 12,227 | 42 | | 1.53 | 1(Ref.) |
|  | Yes | 26,689 | 225 | 2.52 | 1.03 (0.76-1.40) | 18,625 | 121 | | 2.80 | 1.33 (0.84-2.09) |
| ***Trastuzumab*** | No | 36,766 | 269 | 2.18 | 1(Ref.) | 25,672 | 136 | | 2.29 | 1(Ref.) |
|  | Yes | 7,899 | 59 | 2.40 | 0.94 (0.70-1.25) | 5,180 | 27 | | 2.40 | 0.88 (0.58-1.35) |
| ***Endocrine treatment*** | No | 14,849 | 112 | 2.28 | 1(Ref.) | 10,226 | 48 | | 2.05 | 1(Ref.) |
|  | Tamoxifen | 10,631 | 60 | 1.66 | 0.92 (0.67-1.26) | 7,482 | 36 | | 2.06 | 1.34 (0.87-2.09) |
|  | AIs | 18,370 | 152 | 2.53 | 1.02 (0.79-1.30) | 12,562 | 76 | | 2.66 | 1.20 (0.83-1.73) |
|  | Both | 815 | 4 | 1.44 | 0.62 (0.23-1.67) | 582 | 3 | | 2.26 | 1.13 (0.35-3.64) |
| ***Radiation treatment*** | No | 12,263 | 93 | 2.28 | 1(Ref.) | 8,338 | 50 | | 2.53 | 1(Ref.) |
|  | Yes | 32,402 | 235 | 2.19 | 0.99 (0.78-1.27) | 22,514 | 113 | | 2.22 | 0.90 (0.64-1.26) |
| ***Age≥66*** | | Subjects (N) | Case (n) | IR | sHR (95% CI) | Subjects (N) | Case (n) | IR | | sHR (95% CI) |
| ***Anthracycline*** | No | 8,983 | 225 | 8.23 | 1(Ref.) | 5,640 | 109 | | 8.81 | 1(Ref.) |
|  | Yes | 3,161 | 76 | 7.59 | 1.25 (0.90-1.75) | 2,111 | 39 | | 8.40 | 1.23 (0.77-1.97) |
| ***Taxane*** | No | 7,445 | 198 | 8.65 | 1(Ref.) | 4,766 | 99 | | 9.53 | 1(Ref.) |
|  | Yes | 4,699 | 103 | 7.12 | 0.79 (0.58-1.06) | 2,985 | 49 | | 7.38 | 0.75(0.49-1.15) |
| ***Trastuzumab*** | No | 10,893 | 269 | 7.95 | 1(Ref.) | 7,001 | 133 | | 8.56 | 1(Ref.) |
|  | Yes | 1,251 | 32 | 9.09 | 1.08 (0.74-1.58) | 750 | 15 | | 10.13 | 1.04 (0.60-1.82) |
| ***Endocrine treatment*** | No | 3,700 | 95 | 8.49 | 1(Ref.) | 2,314 | 43 | | 8.54 | 1(Ref.) |
|  | Tamoxifen | 2,057 | 53 | 7.88 | 0.92 (0.65-1.29) | 1,406 | 30 | | 9.45 | 1.17 (0.73-1.88) |
|  | AIs | 6,126 | 149 | 8.00 | 0.99 (0.76-1.28) | 3,859 | 73 | | 8.66 | 1.11 (0.76-1.63) |
|  | Both | 261 | 4 | 4.84 | 0.54 (0.20-1.46) | 172 | 2 | | 5.32 | 0.60 (0.14-2.46) |
| ***Radiation treatment*** | No | 5,748 | 155 | 8.73 | 1(Ref.) | 3,671 | 81 | | 9.90 | 1(Ref.) |
|  | Yes | 6,396 | 146 | 7.45 | 1.01 (0.80-1.28) | 4,080 | 67 | | 7.58 | 0.89 (0.64-1.25) |

IR, incidence rate (described as per 1,000 person-years); sHR, sub-distribution hazard ratio; CI, confidence interval; AIs, aromatase inhibitors.

Hazard ratios were adjusted for income, area of residence, hypertension, diabetes mellitus, dyslipidemia, coronary heart disease, congestive heart failure, chronic kidney disease, chronic obstructive pulmonary disease, and use of anthracyclines, taxane, trastuzumab, endocrine therapy, and radiation therapy.

**Supplemental Table 5.** Adjusted sub-distribution hazard ratios for developing atrial fibrillation in breast cancer surgery survivors compared to the noncancer general population stratified by selected factors

|  | | | All ages | | | | Aged 18-50 years | | | |
| --- | --- | --- | --- | --- | --- | --- | --- | --- | --- | --- |
|  |  |  | Subjects (N) | Case (n) | IR per  1,000 PYs | sHR (95% CI) | Subjects (N) | Case (n) | IR per  1,000 PYs | sHR (95% CI) |
| ***Income*** | *High* | Noncancer | 425,672 | 3,799 | 1.69 | 1(Ref.) | 200,799 | 551 | 0.51 | 1(Ref.) |
|  |  | Breast cancer | 87,725 | 884 | 1.96 | 1.07 (0.99-1.15) | 41,590 | 164 | 0.75 | 1.36 (1.14-1.62) |
|  | *Low* | Noncancer | 140,488 | 1,472 | 2.00 | 1(Ref.) | 62,961 | 224 | 0.66 | 1(Ref.) |
|  |  | Breast cancer | 25,507 | 282 | 2.19 | 1.05 (0.92-1.19) | 11,162 | 58 | 1.00 | 1.39 (1.04-1.86) |
|  | P for interaction | | 0.809 | | | | 0.891 | | | |
| ***Residence*** | *Urban* | Noncancer | 265,234 | 2,204 | 1.57 | 1(Ref.) | 124,104 | 325 | 0.48 | 1(Ref.) |
|  |  | Breast cancer | 56,703 | 575 | 1.96 | 1.12 (1.02-1.23) | 25,930 | 95 | 0.69 | 1.34 (1.07-1.69) |
|  | *Rural* | Noncancer | 300,926 | 3,067 | 1.94 | 1(Ref.) | 139,656 | 450 | 0.60 | 1(Ref.) |
|  |  | Breast cancer | 56,529 | 591 | 2.06 | 1.02 (0.93-1.11) | 26,822 | 127 | 0.91 | 1.39 (1.14-1.69) |
|  | P for interaction | | 0.138 | | | | 0.827 | | | |
| ***Hypertension*** | *No* | Noncancer | 443,529 | 2,348 | 1.00 | 1(Ref.) | 245,810 | 618 | 0.47 | 1(Ref.) |
|  |  | Breast cancer | 84,453 | 491 | 1.12 | 1.14 (1.03-1.25) | 47,906 | 167 | 0.66 | 1.37 (1.16-1.63) |
|  | *Yes* | Noncancer | 122,631 | 2,923 | 4.62 | 1(Ref.) | 17,950 | 157 | 1.60 | 1(Ref.) |
|  |  | Breast cancer | 28,779 | 675 | 4.72 | 1.02 (0.93-1.11) | 4,846 | 55 | 2.18 | 1.35 (0.99-1.84) |
|  | P for interaction | | 0.087 | | | | 0.939 | | | |
| ***DM*** | *No* | Noncancer | 526,404 | 4,318 | 1.55 | 1(Ref.) | 258,030 | 729 | 0.52 | 1(Ref.) |
|  |  | Breast cancer | 103,205 | 920 | 1.73 | 1.08 (1.01-1.16) | 51,225 | 208 | 0.77 | 1.37 (1.18-1.61) |
|  | *Yes* | Noncancer | 39,756 | 953 | 4.79 | 1(Ref.) | 5,730 | 46 | 1.52 | 1(Ref.) |
|  |  | Breast cancer | 10,027 | 246 | 5.06 | 1.00 (0.87-1.15) | 1,527 | 14 | 1.81 | 1.28 (0.71-2.34) |
|  | P for interaction | | 0.302 | | | | 0.829 | | | |
| ***Dyslipidemia*** | *No* | Noncancer | 465,426 | 3,496 | 1.41 | 1(Ref.) | 250,354 | 684 | 0.51 | 1(Ref.) |
|  |  | Breast cancer | 90,164 | 751 | 1.60 | 1.10 (1.01-1.19) | 49,231 | 200 | 0.77 | 1.42 (1.21-1.66) |
|  | *Yes* | Noncancer | 100,734 | 1,775 | 3.55 | 1(Ref.) | 13,406 | 91 | 1.30 | 1(Ref.) |
|  |  | Breast cancer | 23,068 | 415 | 3.69 | 1.01 (0.91-1.12) | 3,521 | 22 | 1.22 | 1.04 (0.65-1.65) |
|  | P for interaction | | 0.224 | | | | 0.211 | | | |
| ***CHD*** | *No* | Noncancer | 532,649 | 4,122 | 1.46 | 1(Ref.) | 258,706 | 706 | 0.51 | 1(Ref.) |
|  |  | Breast cancer | 104,500 | 906 | 1.68 | 1.10 (1.03-1.19) | 50,936 | 200 | 0.75 | 1.41 (1.20-1.65) |
|  | *Yes* | Noncancer | 33,511 | 1,149 | 6.75 | 1(Ref.) | 5,054 | 69 | 2.53 | 1(Ref.) |
|  |  | Breast cancer | 8,732 | 260 | 6.06 | 0.94 (0.82-1.08) | 1,816 | 22 | 2.39 | 1.05 (0.65-1.71) |
|  | P for interaction | | **0.042** | | | | 0.261 | | | |
| ***CHF*** | *No* | Noncancer | 558,490 | 4,854 | 1.64 | 1(Ref.) | 262,751 | 752 | 0.53 | 1(Ref.) |
|  |  | Breast cancer | 110,186 | 1,057 | 1.86 | 1.07 (1,10-1.44) | 51,920 | 212 | 0.78 | 1.39 (1.19-1.63) |
|  | *Yes* | Noncancer | 7,670 | 417 | 12.04 | 1(Ref.) | 1,009 | 23 | 4.76 | 1(Ref.) |
|  |  | Breast cancer | 3,046 | 109 | 8.63 | 1.01 (0.82-1.25) | 832 | 10 | 2.90 | 0.91 (0.43-1.92) |
|  | P for interaction | | 0.624 | | | | 0.276 | | | |
| ***CKD*** | *No* | Noncancer | 561,716 | 5,075 | 1.71 | 1(Ref.) | 262,762 | 760 | 0.54 | 1(Ref.) |
|  |  | Breast cancer | 111,947 | 1,107 | 1.92 | 1.06 (1.00-1.14) | 52,428 | 218 | 0.79 | 1.38 (1.18-1.61) |
|  | *Yes* | Noncancer | 4,444 | 196 | 9.66 | 1(Ref.) | 998 | 15 | 2.98 | 1(Ref.) |
|  |  | Breast cancer | 1,285 | 59 | 10.57 | 1.07 (0.80-1.43) | 324 | 4 | 2.60 | 0.92 (0.30-2.77) |
|  | P for interaction | | 0.975 | | | | 0.476 | | | |
| ***COPD*** | *No* | Noncancer | 512,643 | 4,290 | 1.58 | 1(Ref.) | 247,404 | 710 | 0.53 | 1(Ref.) |
|  |  | Breast cancer | 92,534 | 842 | 1.77 | 1.11 (1.03-1.19) | 45,370 | 179 | 0.75 | 1.35 (1.14-1.59) |
|  | *Yes* | Noncancer | 53,517 | 981 | 3.53 | 1(Ref.) | 16,356 | 65 | 0.74 | 1(Ref.) |
|  |  | Breast cancer | 20,698 | 324 | 3.05 | 0.96 (0.85-1.09) | 7,382 | 43 | 1.09 | 1.51 (1.02-2.22) |
|  | P for interaction | | **0.055** | | | | 0.596 | | | |

IR, incidence rate; PYs, person-years; HR, hazard ratio; sHR, sub-distribution hazard ratio; CI, confidence interval; HTN, hypertension; DM, diabetes mellitus; DL, dyslipidemia; CHD, coronary heart disease; CHF, congestive heart failure; CKD, chronic kidney disease; COPD; chronic obstructive pulmonary disease.

Each model was adjusted for age, income, area of residence, hypertension, diabetes mellitus, dyslipidemia, coronary heart disease, congestive heart failure, chronic kidney disease, and chronic obstructive pulmonary disease, but not for the covariate used in stratified analysis.

**Supplemental Table 6.** Adjusted sub-distribution hazard ratios for developing atrial fibrillation in breast cancer surgery survivors compared to the noncancer general population by age categories: A s**ensitivity analysis after including person-time within the first year of follow-up**

|  | | | sHR (95% CI) |
| --- | --- | --- | --- |
| ***Age group*** | ***All ages*** | Noncancer | 1(Ref.) |
|  |  | Breast cancer | 1.60(1.52-1.69) |
|  | ***18-39*** | Noncancer | 1(Ref.) |
|  |  | Breast cancer | 6.84(5.18-9.03) |
|  | ***40-50*** | Noncancer | 1(Ref.) |
|  |  | Breast cancer | 2.81(2.48-3.19) |
|  | ***51-65*** | Noncancer | 1(Ref.) |
|  |  | Breast cancer | 1.71(1.58-1.85) |
|  | ***≥66*** | Noncancer | 1(Ref.) |
|  |  | Breast cancer | 1.01(0.92-1.11) |
| *P* for interaction | | | <0.001 |

sHR, sub-distribution hazard ratio; CI, confidence interval;

Adjusted for age, income, area of residence, hypertension, diabetes mellitus, dyslipidemia, coronary heart disease, congestive heart failure, chronic kidney disease, and chronic obstructive pulmonary disease

**Supplemental Table 7.** Adjusted sub-distribution hazard ratios for developing atrial fibrillation by cancer treatment type among breast cancer surgery survivors by age categories: A s**ensitivity analysis after including person-time within the first year of follow-up**

|  | | All ages | Age 18-39 | Age 40-50 | Age 51-65 | Age ≥66 |
| --- | --- | --- | --- | --- | --- | --- |
|  |  | sHR (95% CI) | sHR (95% CI) | sHR (95% CI) | sHR (95% CI) | sHR (95% CI) |
| ***Anthracyclines*** | No | 1(Ref.) | 1(Ref.) | 1(Ref.) | 1(Ref.) | 1(Ref.) |
|  | Yes | 1.72(1.48-2.00) | 2.24 (1.29-4.51) | 1.86 (1.30-2.65) | 1.81 (1.44-2.28) | 1.51 (1.15-1.98) |
| ***Taxane*** | No | 1(Ref.) | 1(Ref.) | 1(Ref.) | 1(Ref.) | 1(Ref.) |
|  | Yes | 0.81(0.70-0.94) | 0.68 (0.39-1.18) | 0.90 (0.63-1.28) | 0.79 (0.63-1.00) | 0.90 (0.69-1.16) |
| ***Trastuzumab*** | No | 1(Ref.) | 1(Ref.) | 1(Ref.) | 1(Ref.) | 1(Ref.) |
|  | Yes | 0.97(0.86-1.10) | 0.92 (0.58-1.43) | 1.04 (0.80-1.35) | 0.91 (0.76-1.09) | 1.14 (0.87-1.49) |
| ***Endocrine therapy*** | No | 1(Ref.) | 1(Ref.) | 1(Ref.) | 1(Ref.) | 1(Ref.) |
|  | Tamoxifen | 0.89(0.79-1.00) | 0.90 (0.63-1.27) | 0.92 (0.73-1.16) | 0.83 (0.67-1.03) | 0.88 (0.67-1.16) |
|  | AIs | 0.98(0.88-1.10) | 1.97 (0.27-14.2) | 0.88 (0.51-1.53) | 0.92 (0.79-1.08) | 1.13 (0.93-1.38) |
|  | Both | 0.89(0.62-1.29) | N/A | 2.62 (1.06-6.44) | 0.82 (0.41-1.26) | 0.91 (0.50-1.63) |
| ***Radiation treatment*** | No | 1(Ref.) | 1(Ref.) | 1(Ref.) | 1(Ref.) | 1(Ref.) |
|  | Yes | 0.89(0.81-0.98) | 1.04 (0.69-1.54) | 0.78 (0.63-0.96) | 0.93 (0.79-1.08) | 0.96 (0.81-1.14) |

sHR, sub-distribution hazard ratio; CI, confidence interval; AIs, aromatase inhibitors.

Adjusted for age, income, area of residence, hypertension, diabetes mellitus, dyslipidemia, coronary heart disease, congestive heart failure, chronic kidney disease, and chronic obstructive pulmonary disease, history of anthracycline, taxane, trastuzumab, endocrine treatment and radiation treatment

**Supplemental Table 8.** Adjusted sub-distribution hazard ratios for developing atrial fibrillation in breast cancer surgery survivors compared to the noncancer general population by age categories: **A subset analysis based on data comprising participants in the general health screening examination**

| **Age group** | | Subjects (N) | Case (n) | IR per  1,000 PYs | Model 1 (Crude)  HR (95% CI) | Model 2  sHR (95% CI) | Model 3  sHR (95% CI) | Model 4  sHR (95% CI) |
| --- | --- | --- | --- | --- | --- | --- | --- | --- |
| ***All ages*** | Noncancer | 292,468 | 2,535 | 1.70 | 1(Ref.) | 1(Ref.) | 1(Ref.) | 1(Ref.) |
|  | Breast cancer | 72,560 | 725 | 1.98 | 1.17 (1.08-1.27) | 1.18 (1.09-1.28) | 1.11 (1.02-1.21) | 1.10(1.01-1.19) |
| ***18-39*** | Noncancer | 15,347 | 16 | 0.20 | 1(Ref.) | 1(Ref.) | 1(Ref.) | 1(Ref.) |
|  | Breast cancer | 3,608 | 13 | 0.71 | 3.59 (1.73-7.45) | 3.57 (1.72-7.41) | 3.40 (1.64-7.08) | 3.44 (1.66-7.16) |
| ***40-50*** | Noncancer | 102,950 | 302 | 0.57 | 1(Ref.) | 1(Ref.) | 1(Ref.) | 1(Ref.) |
|  | Breast cancer | 26,232 | 96 | 0.71 | 1.26 (1.00-1.58) | 1.27 (1.01-1.60) | 1.21 (0.96-1.52) | 1.22 (0.97-1.53) |
| ***51-65*** | Noncancer | 138,354 | 1,146 | 1.62 | 1(Ref.) | 1(Ref.) | 1(Ref.) | 1(Ref.) |
|  | Breast cancer | 33,831 | 352 | 2.07 | 1.28 (1.13-1.44) | 1.30 (1.15-1.46) | 1.22 (1.08-1.38) | 1.21 (1.07-1.37) |
| ***≥66*** | Noncancer | 35,817 | 1,071 | 6.17 | 1(Ref.) | 1(Ref.) | 1(Ref.) | 1(Ref.) |
|  | Breast cancer | 8,889 | 264 | 6.21 | 1.01 (0.88-1.15) | 1.00 (0.88-1.15) | 0.94 (0.82-1.08) | 0.92 (0.80-1.05) |
| P for interaction | |  |  |  | 0.001 | 0.001 | 0.001 | <0.001 |

IR, incidence rate; PYs, person-years; HR, hazard ratio; sHR, sub-distribution hazard ratio; CI, confidence interval.

Model 2 was adjusted for age, income, and area of residence. Model 3 was adjusted for age, income, area of residence, hypertension, diabetes mellitus, dyslipidemia, coronary heart disease, congestive heart failure, chronic kidney disease, and chronic obstructive pulmonary disease. Model 4 was additionally adjusted for body mass index, smoking status, alcohol consumption and regular physical activity from Model 3.

**Supplemental Table 9.** Adjusted sub-distribution hazard ratios for developing atrial fibrillation by cancer treatment type among breast cancer surgery survivors: **A subset analysis based on data comprising participants in the general health screening examination**

| **Treatment type** | | Subjects (N) | Case (n) | IR | Model 1 (Crude)  HR (95% CI) | Model 2  sHR (95% CI) | Model 3  sHR (95% CI) | Model 4  sHR (95% CI) | Model 5  sHR (95% CI) |
| --- | --- | --- | --- | --- | --- | --- | --- | --- | --- |
| ***Anthracycline*** | No | 36,492 | 369 | 2.03 | 1(Ref.) | 1(Ref.) | 1(Ref.) | 1(Ref.) | 1(Ref.) |
|  | Yes | 36,068 | 356 | 1.94 | 0.95 (0.82-1.10) | 1.41 (1.21-1.64) | 1.37 (1.18-1.60) | 1.57 (1.21-2.03) | 1.55 (1.20-2.01) |
| ***Taxane*** | No | 31,595 | 325 | 2.06 | 1(Ref.) | 1(Ref.) | 1(Ref.) | 1(Ref.) | 1(Ref.) |
|  | Yes | 40,965 | 400 | 1.93 | 0.94 (0.81-1.08) | 1.24 (1.06-1.44) | 1.19 (1.03-1.39) | 0.83 (0.65-1.08) | 0.83 (0.64-1.07) |
| ***Trastuzumab*** | No | 61,922 | 617 | 1.96 | 1(Ref.) | 1(Ref.) | 1(Ref.) | 1(Ref.) | 1(Ref.) |
|  | Yes | 10,638 | 108 | 2.09 | 1.07 (0.87-1.32) | 1.17 (0.95-1.43) | 1.11 (0.90-1.36) | 0.95 (0.76-1.19) | 0.95 (0.77-1.19) |
| ***Endocrine treatment*** | No | 22,012 | 239 | 2.21 | 1(Ref.) | 1(Ref.) | 1(Ref.) | 1(Ref.) | 1(Ref.) |
|  | Tamoxifen | 30,313 | 173 | 1.10 | 0.50(0.41-0.61) | 0.78 (0.64-0.96) | 0.78 (0.64-0.95) | 0.81 (0.66-0.99) | 0.81 (0.66-0.99) |
|  | AIs | 19,317 | 302 | 3.14 | 1.42 (1.20-1.69) | 0.98 (0.82-1.16) | 0.97 (0.82-1.15) | 0.98 (0.82-1.17) | 0.96 (0.81-1.15) |
|  | Both | 918 | 11 | 2.39 | 1.08 (0.59-1.98) | 0.78 (0.43-1.43) | 0.73 (0.40-1.33) | 0.75 (0.41-1.37) | 0.74 (0.40-1.35) |
| ***Radiation treatment*** | No | 20,973 | 257 | 2.47 | 1(Ref.) | 1(Ref.) | 1(Ref.) | 1(Ref.) | 1(Ref.) |
|  | Yes | 51,587 | 468 | 1.79 | 0.73 (0.62-0.85) | 0.97 (0.83-1.14) | 0.99 (0.85-1.16) | 0.96 (0.82-1.12) | 0.95 (0.81-1.11) |

IR, incidence rate (described as per 1,000 person-years); HR, hazard ratio; sHR, sub-distribution hazard ratio; CI, confidence interval; AIs, aromatase inhibitors.

Model 2 was adjusted for age, income, and area of residence. Model 3 was adjusted for age, income, area of residence, hypertension, diabetes mellitus, dyslipidemia, coronary heart disease, congestive heart failure, chronic kidney disease, and chronic obstructive pulmonary disease. Model 4 was additionally adjusted for history of anthracycline, taxane, trastuzumab, endocrine treatment and radiation treatment from Model 3. Model 5 was additionally adjusted for body mass index, smoking status, alcohol consumption and regular physical activity from Model 4.
